# Serological testing in addition to PCR screening for the re-opening of American colleges and universities: potential for cost-savings without compromising pandemic mitigation

**DOI:** 10.1101/2020.10.04.20206680

**Authors:** Youngji Jo, Ruby Singh, Gabriella Rao, Sandro Galea, Brooke Nichols

## Abstract

**Importance:** The addition of a serological testing could reduce the overall testing costs of a PCR-based SARS-CoV-2 testing reopening plan for colleges/universities in the United States, without compromising the efficacy of the testing plan.

**Objectives:** To determine whether a college/university reopening SARS-CoV-2 testing plan that includes serological testing can be cost-saving compared to a PCR-only testing.

**Design, Setting, and Participants:** We assessed costs of serological testing in addition to PCR testing under various scenarios of university sizes (2000, 10,000, and 40,000) and epidemic conditions (initial antibody prevalence 2.5-15%; cumulative SARS-CoV-2 incidence during the school year 5-30%) of SARS-CoV-2 in the United States. We estimated total testing costs and relative percentage of cost-savings of different screening (i.e. targeted/ universal) and testing (i.e. in-sourcing/out-sourcing) scenarios between September 2020-May 2021.

**Main Outcomes and Measures:** Testing costs of serological testing and PCR testing, Relative percentage of cost saving by including serology testing in addition to PCR testing.

**Results:** Including baseline serology testing alongside routine regular PCR testing can reduce total test volumes and related costs throughout the school year. While the total testing cost is likely much lower if regular PCR testing is insourced compared to outsourced ($5 million vs $34 million for university size 10,000), including serologic testing could achieve the up to 20% cost-savings relative to PCR testing alone. The insourcing of serological testing when PCR testing is insourced can achieve greater cost-savings under high initial antibody prevalence (>5%) and cumulative incidence throughout the school year (>10%) at medium and large sized universities. If PCR testing is outsourced, however, the inclusion of serological testing becomes always preferred in most university sizes and epidemic conditions.

**Conclusions and Relevance:** While regular PCR testing alone is the preferred strategy for containing epidemics, including serology testing may help achieve cost-savings if outbreaks are anticipated, or if baseline seropositivity is high.

**Key Points (96/100):** *Question:* Can the addition of a serological testing reduce the overall testing costs of a PCR-based SARS-CoV-2 testing reopening plan for universities in the United States?

*Findings:* This costing study suggested that inclusion of serological testing in addition to outsourced PCR testing as part of a university re-opening strategy could achieve cost savings of up to 20%. The amount of savings, or additional costs, is dependent on insourcing or outsourcing of testing, epidemic conditions and university size.

*Meaning:* The relative cost-savings depend strongly on whether PCR and/or serology are being insourced or outsourced, university sizes and cumulative incidence.

## Introduction

In the fall of 2020, colleges and universities throughout the United States started to open their doors to in-person education amid the COVID-19 pandemic. Many adopted a testing-based opening strategy as a piece of their mitigation strategies to contain outbreaks of SARS-CoV-2 on college campuses, enabling the restart of in-person education. In order for this testing strategy to be successful, reverse transcription polymerase chain reaction testing (RT-PCR, or PCR for short) must be conducted at very frequent intervals, as many as once every two days for those in high-contact with others on campus.^1^ However, this frequent PCR testing may be cost-prohibitive for the majority of American colleges and universities suggesting that ways to reduce costs without compromising the testing strategy should be considered.

The addition of serological testing to the PCR testing algorithm may be one such way to reduce overall costs to a college re-opening plan. While antibodies may only be detectable through currently available tests for a limited duration^2^ it is thought that immunity to the virus may persist beyond the period of detectable antibodies through a T-cell mediated response.^3^ Therefore, someone who tests antibody positive with a serological test may be exempt from repeat PCR testing during the school year, resulting in cost savings to the institution or payer, and reduced burden on members of the university community.

Therefore, we assessed costs of including serological testing in addition to regular and repeated PCR testing under various university sizes and epidemic conditions in the United States. We explored different screening (i.e. targeted/ universal) and testing (i.e. in-sourcing/out-sourcing of PCR testing) strategies to determine in what instances serological testing could be a cost-saving addition to a PCR-only testing strategy.

## Methods

We developed a model in Microsoft Excel to calculate the total number of PCR tests required, by university community size, for an effective testing-based campus opening strategy over a 32-week (two semesters) time horizon (September 2020-May 2021). For our PCR testing algorithm, we followed the PCR testing protocol outlined by Boston University. We categorized the students, faculty, and staff into the four groups (category 1-4) based on the severity of potential exposure to COVID-19, apportioned to 40%, 20%, 20%, and 20% of the target population respectively. The frequencies of testing were based on accepted COVID-19 transmission models taking into account the specifics of spread within the university community^4^ The categories included:

- Category 1 (PCR tests twice per week): Commuting students, staff, and faculty who interact with residential students for significant periods of time either in classes or other activities or who otherwise spend many hours on campus in close-contact activities like athletics, performing arts, or in some research and off-campus educational environments
- Category 2 (PCR tests once per week): Commuting students residing off-campus attending in-person classes, but with little contact with residential students
- Category 3 (PCR tests once per month): Commuting employees whose job duties require very limited contact with students and who can control their contact with other employees so as to limit interactions to small groups of individuals with appropriate work environment protocols in place and minimal contact hours
- Category 4 (No PCR tests): Students, faculty, and staff who engage only in virtual learning, working and other activities and events and who do not commute to campus

For the scenario that includes serological testing, we assume that individuals in categories 1-3 will receive an antibody test upon arrival on campus alongside their initial PCR test (Figure 1). If the individual is antibody positive, we assume that they will not be tested again for the remainder of the year. If they are PCR positive and antibody negative upon arrival to campus, an antibody test will be re-done 30-days post symptom onset. If the antibody test remains negative, then the individual will return to the PCR testing algorithm. At the beginning of the second semester, everyone who has never tested antibody positive will again have a serological test and a PCR test upon arrival. This is to enable the university to detect anyone who may have been infected during the break.

**Figure 1.**
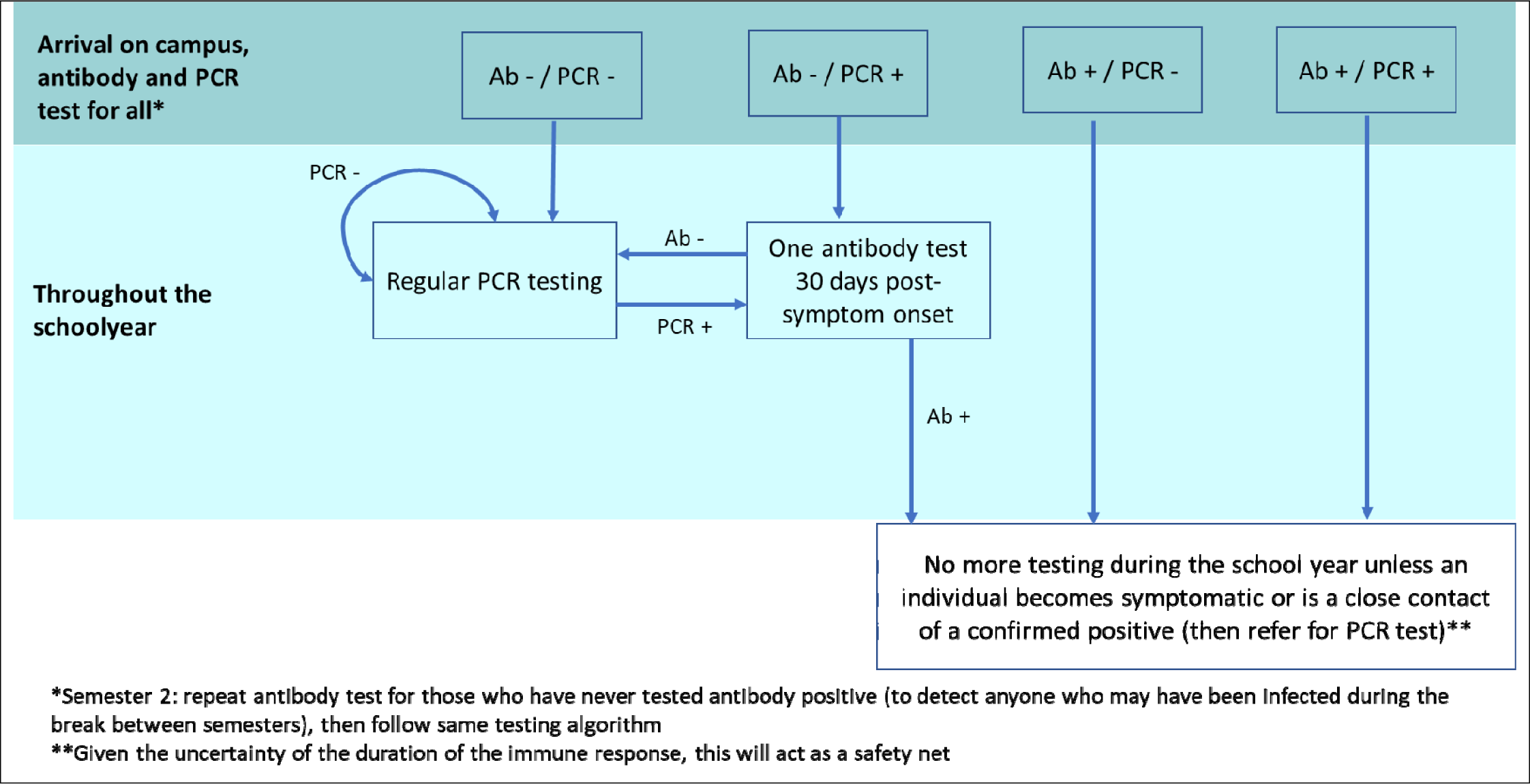
Proposed testing algorithm that includes serological testing (Antibody, Ab) in addition to regular PCR testing.

We ranged the initial prevalence of antibodies at the beginning of the fall semester between 2.5%-15%, and ranged the cumulative proportion of the university community infected and PCR-confirmed during the school year between 5-30%. We assumed that the immune response remains throughout the school year (32 weeks).^5,6^ The total number of positive antibody tests is a sum of the baseline prevalence plus the number of incident PCR-positive tests during the school year. Based on the estimates, we calculated the total number of PCR and serological tests required (assuming serological test sensitivity as 0.915) during the school year for each testing category respectively, which are combined with relevant testing frequency and unit costs per testing ($100 if PCR outsourced and $50 if serological testing outsourced^7^) for each category respectively. Together with the serological testing costs ($15 if PCR insourced and $7.5 if serological testing insourced^8^), we evaluated total testing costs by multiple scenarios of in-sourcing and out-sourcing both PCR and antibody testing. For insourcing strategies, we additionally included capital and recurrent costs of setting up and running the in-house laboratory infrastructure. (Table 1)

**Table 1.** Key input parameters.

| University size | Base case | Low | High | Source |
| --- | --- | --- | --- | --- |
| Total population size | 10,000 | 2,000 | 40,000 | 9 |
| Track 1 (PCR 2 per week) | 40% | NA | NA | 4 |
| Track 2 (PCR 1 per week) | 20% | NA | NA |  |
| Track 3 (PCR 1 per month) | 20% | NA | NA |  |
| Track 4 (No testing) | 20% | NA | NA |  |
| Number of weeks | 32 | NA | NA |  |
| Epidemic condition |  |  |  |  |
| Prevalence of antibodies on arrival | 5% | 2.5% | 15% | 1,10,12,13 |
| Proportion of people expected to test PCR pos | 10% | 5% | 30% |  |
| Abbott Architect IgG | 0.915 | 0.8 | 0.98 | 24 |
| Cost |  |  |  |  |
| Capital cost for lab setting for PCR test | \$150,000 | \$100,000 | \$200,000 | 7 |
| Recurrent cost for PCR lab operation per year | \$20,000 | \$15,000 | \$25,000 | |
| Cost per Rt-PCR test (In-sourcing) | \$15 | \$10 | \$20 | |
| Cost per Rt-PCR test (Out-sourcing) | \$100 | \$80 | \$200 | |
| Capital cost for serological testing (Abbott architect i2000SR instrument) | \$107,500 | \$86,000 | \$129,000 | 8 |
| Re-training option per year | \$2,955 | \$2,364 | \$3,546 | |
| Recurrent cost for Architect i2000SR per year | \$13,200 | \$10,560 | \$15,840 | |
| Cost per Abbott serological test (In-sourcing) | \$7.5 | \$5 | \$10 | |
| Cost per Abbott serological test (Out-sourcing) | \$50 | \$40 | \$150 | |

We considered three different size university settings with a target population of 2,000, 10,000, and 40,000 community members^9^ and base case of the initial detectable antibody prevalence of 5% (varied from 2.5% to 15%) on September 2020 and the cumulative PCR-positive university community members as 10% (varied from 5 to 30%) by May, 2021. As additional scenario analyses, we also considered a targeted serological testing approach, where only category 1 university community members receive a serological test while category 1-3 members still receive the regular PCR testing, given the greatest likelihood of cost-savings in this category due to multiple PCR tests per week. Outputs were estimated as total costs and relative percentage of cost saving of the complete testing strategy by arm including serological testing in addition to PCR testing alone for the respective screening and testing strategies over the two semesters (32 weeks). To test the robustness of our cost-saving estimates, we performed a number of sensitivity analyses (one-way and three-way sensitivity analyses) based on the uncertainty estimates of each parameter value.

## Results

Total testing cost and relative percentage of cost saving differ by university size, initial antibody prevalence, cumulative PCR-confirmed incidence across the school-year, as well as by screening/testing strategies (Table 2). For a medium sized university setting (10,000 students) assuming a 5% initial prevalence of antibodies and 10% cumulative incidence of SARS-CoV-2 during September 2020-May 2021, universal screening (80% of the university community) of PCR testing, without the addition of serological testing, costs approximately $34.4 million when outsourced and $5.3 million when insourced. With the addition of serological testing in this scenario, the total cost drops to $32 million when PCR is outsourced. When PCR is insourced, the total cost slightly drops to $5.2 million with insourced serological testing but slightly increases to $5.8 million with outsourced serological testing. Targeted screening of Category 1 for serological testing (40% of target population) will decrease costs from $800,000 to $400,000 if serology testing is outsourced and $244,000 to $184,000, if serologic testing is insourced, which results in a <8% decrease in total testing costs compared to universal screening across categories. This is due to the fact that regular PCR testing costs are substantial (more than 90% out of total testing costs) compared to serological testing costs.

**Table 2.** Total costs and cost saving by including serological testing in addition to PCR testing under initial antibody prevalence of 5%, cumulative SARS-CoV-2 incidence during the school year of 10% (uncertainty ranges are based on varying initial prevalence 2.5-15%; and cumulative prevalence 5-30%) with universal antibody screening at the beginning of each semester at a medium sized university size (10,000)

| Testing strategies | | Cost (\$USD, thousands) | | | |
| --- | --- | --- | --- | --- | --- |
| PCR | Serology | PCR testing | Serological testing | Total testing | Cost savings |
| PCR insourcing | No testing | \$5,330 | NA | \$5,330 | ref |
| | Insourcing | \$4,814<br>(\$4,122 - \$4,987) | \$244 | \$5,228<br>(\$4,536 - \$5,401) | \$102<br>(-\$71, \$794) |
| | Outsourcing | \$4,814<br>(\$4,122 - \$4,987) | \$800 | \$5,784<br>(\$5,092 - \$5,957) | -\$454<br>(-\$627, \$238) |
| PCR outsourcing | No testing | \$34,400 | NA | \$34,400 | ref |
| | Insourcing | \$32,094<br>(\$27,483 - \$33,247) | \$244 | \$32,338<br>(\$27,726 - \$33,491) | \$2,062<br>(\$909, \$6,674) |
| | Outsourcing | \$32,094<br>(\$27,483 - \$33,247) | \$800 | \$32,894<br>(\$28,283 - \$34,047) | \$1,506<br>(\$353, \$6,117) |

Under a plausible range of epidemic conditions (initial antibody prevalence 2.5-15% and cumulative PCR-confirmed incidence during the schoolyear of 5-30%), including serological testing in addition to PCR outsourcing testing in most university settings could achieve cost savings (1% to 20% depending on epidemic conditions) (Figure 2). On the other hand, including outsourced serological testing in addition to PCR insourced testing may increase costs up to 12% under the same epidemic conditions. While PCR testing alone is the preferred strategy for epidemic containment, including serological testing (particularly when insourced) may result in cost-savings if an outbreak occurs, or if the baseline antibody prevalence is higher than expected. Overall, targeted screening shows a similar pattern as compared to universal screening but a slightly greater relative percentage of cost saving at medium and large sized universities.

**Figure 2.**
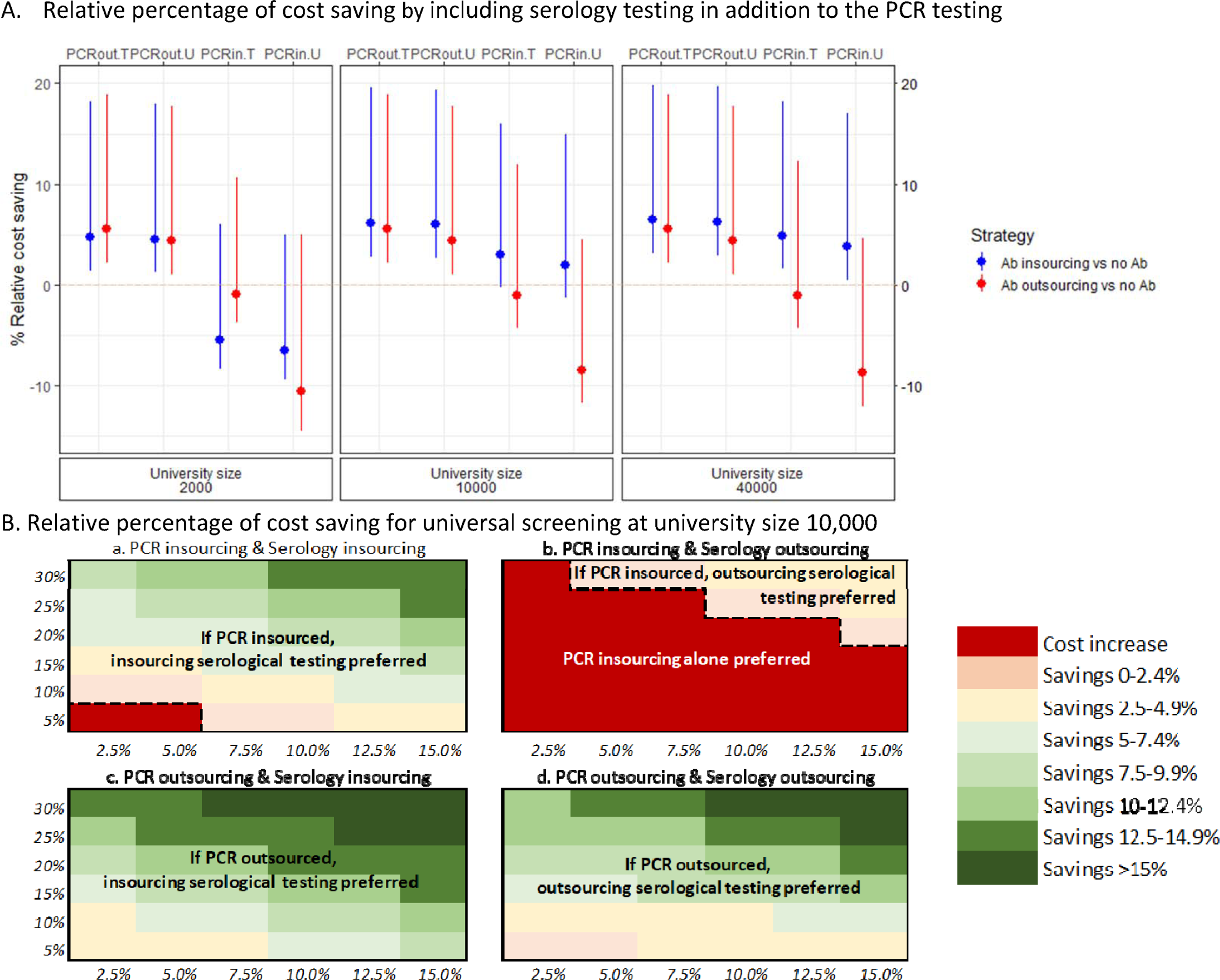
Relative percentage of cost saving by including serology testing in addition to the PCR testing under epidemic condition by university size and screening and testing strategies. In panel A, the base case is (circular point) set as 5% for initial prevalence and 10% for cumulative incidence. The bar represents the extent of relative percentage of cost saving based on the variation of epidemic condition (initial prevalence 2.5-15%; and cumulative incidence 5-30%). Negative estimates indicate cost increase, while positive estimates indicate cost saving, by including serological testing in addition to the PCR testing. For example, for a targetted screening at University size 10000, insourcing serological testing (Blue bar) in addition to PCR outsourcing (“PCRout.T”) will result in 6% (uncertainty range: from 3% under the initial prevalence and cumulative incidence as 2.5% and 5% to 19% under 15% and 30%) cost saving at basecase epidemic condition. On the other hand, for a universal screening at University size 10000, outsourcing serological testing (Red bar) in addition to PCR insourcing (“PCRin.U”) will result 9% cost increase (uncertainty range: from 12% cost increase under 2.5% and 5% to 4% cost saving under 15% and 30%) at basecase epidemic condition. Panel B illustrates heat maps of the relative percentage of cost saving for universal screening at university size 10,000 across a range of the epidemic conditions (initial prevalence 2.5-15%; and cumulative incidence 5-30%). Dark red (cost increase) represents cost saving thresholds (black dashed lines) under the set of epidemic conditions in the given scenario. Targetted screening shows similar pattern for the most scenarios but greater cost saving estimates for the scenario with outsourcing serological testing to PCR insourcing compared to the universal screening.

Our sensitivity analyses (Figure 3) reveal that when including serological testing in addition to regular PCR testing, cost savings is strongly driven by university population size and cumulative incidence. Not surprisingly, cost savings is likely greater for larger universities and when cumulative incidence, as well as initial prevalence, are high. For example, as we varied cumulative incidence between 5% to 30% from the baseline value of 10% under universal screening at a medium sized university, cost savings of including serological testing varied between $1.3 million to $5 million from the baseline value $2 million if additional serological testing is insourced under the condition of PCR outsourcing testing alone. On the other hand, if additional serological testing is outsourced under PCR insourcing testing condition, reducing serological testing cost is the key determinant to reduce total incremental costs relative to PCR testing alone.

**Figure 3.**
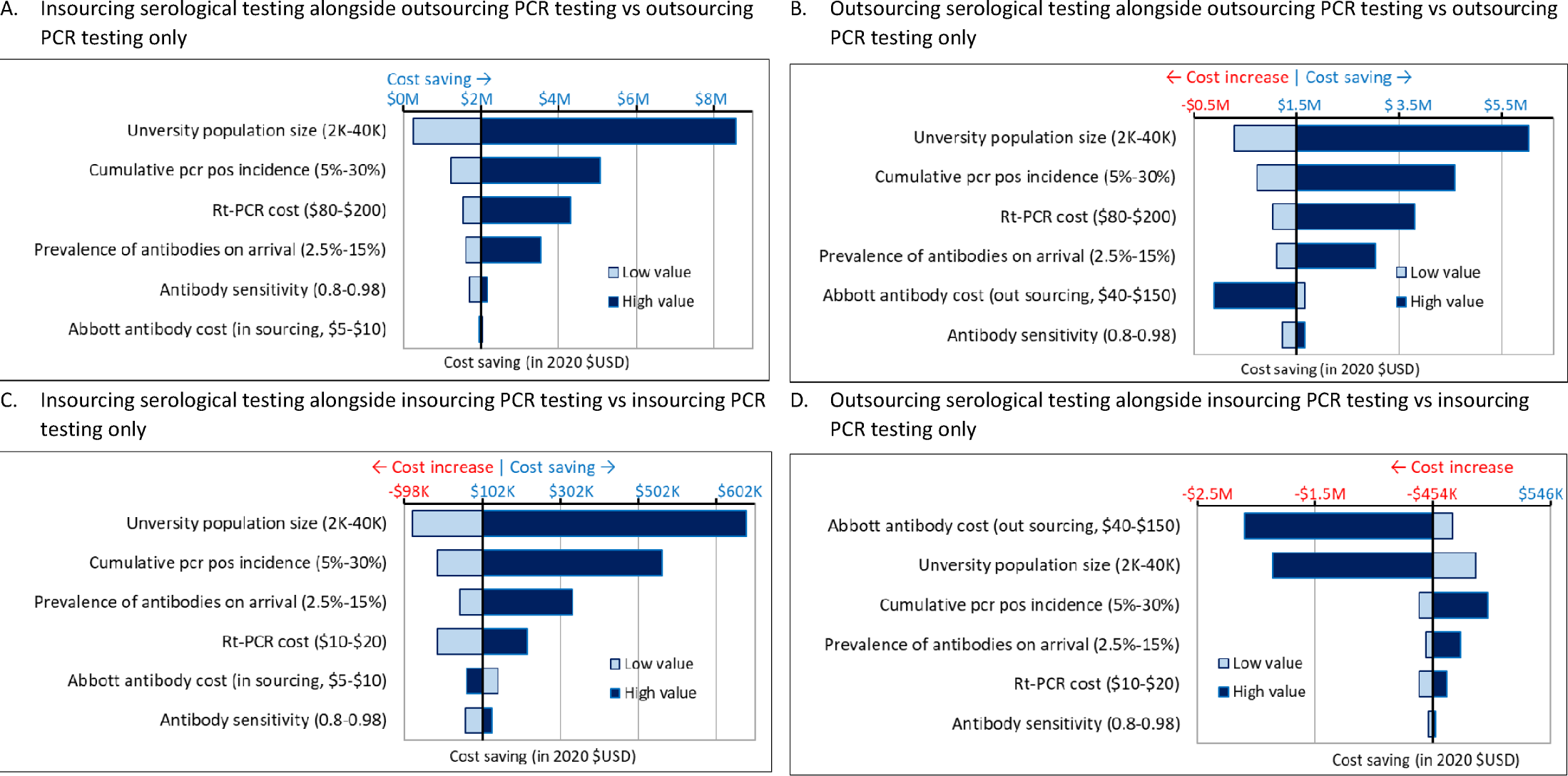
One way sensitivity analyses by including serology testing in addition to the PCR testing for a medium university size (10,000) under 5% initial prevalence and 10% cumulative incidence with universal screening

## Discussion

This analysis explores whether and to what extent PCR and serology screening and testing strategies might be considered cost saving depending on university sizes, epidemic conditions, and screening and testing strategies in the United States. We found that serology testing in addition to regular PCR screening can reduce total test volume and related costs through community members with positive serological test results compared to PCR testing alone. The total testing cost is likely much lower if the regular PCR testing is insourced compared to outsourced ($5 million vs $34 million). While regular and repeated PCR testing is the current preferred strategy for epidemic containment, the inclusion of serological testing may help achieve cost savings if outbreaks occur, or if baseline antibody prevalence is higher than expected, especially in larger sized university communities. In smaller sized universities, however, including serology testing may not be cost-effective if regular PCR testing is highly effective at containing outbreaks.

Rapid and frequent PCR testing is a central pillar of a mitigation strategy to ensure that individuals quickly isolate themselves if positive and hence control outbreaks in the community. Serological testing is important to estimate the prevalence of infections, including those that are asymptomatic. We show that the inclusion of serological testing to a university testing regiment can reduce the required total number and cost of PCR testing. Importantly we found that this was the case only when serological testing was insourced. The outsourcing of serological testing when PCR tests are insourced was rarely cost saving in any scenario compared to a scenario without serological testing. This is because the additional cost required for serological outsourcing testing is greater than the benefit of test volume reduction. The insourcing of serological testing when PCR testing is insourced can achieve greater cost-savings (1%-15%) under high initial antibody prevalence (>5%) and cumulative incidence throughout the school year (>10%) at medium and large sized universities. Despite the high up-front cost of setting up in-house laboratory infrastructure for insourcing of serologic testing, the final resulting cost per test can be much lower if insourced ($7) than outsourced ($50), resulting much lower total serological insourcing testing costs and thus cost saving relative to PCR testing alone. If, however, PCR testing is outsourced, the inclusion of serological testing becomes always preferred in most university sizes and epidemic conditions.

Our study shows that cumulative incidence may be the most important intervenable determinant of cost-savings when including serology testing to PCR testing condition. Several previous studies^1,10,11^ assessed PCR-based SARS-CoV-2 screening strategies – with varying test frequency and sensitivity (and offered interactive tools to project cumulative incidence ^12,13^) – for university settings in the United States. They suggested testing frequency was more strongly associated with cumulative infection than test sensitivity. Studies also found that the spread of COVID-19 is mostly happening during a short period of infectivity (3-5 days) from a subset of people (i.e. an estimated eighty per cent of transmissions are caused by just ten per cent of cases).^14^ Studies to date have not, however, assessed how antibody testing can be incorporated within a university re-opening strategy.

A university-funded testing program may not be feasible for the majority of American colleges and universities. If the individual is responsible for the cost of their own testing, the potential in reduced medical co-payment costs (if insured) through a once or twice-off antibody test may be cost-saving at the individual level.^15^ Additionally, if university community members are responsible for getting themselves tested, they may be less adherent, and as such outbreaks may occur (increasing the cost-savings of serological testing). As the optimal testing frequency likely depends on the baseline prevalence of infection in the group^1^, frequency of regular PCR testing may be reduced (e.g. from two to once a week for category 1) as the percent positive gets lower over time (i.e. below 5% for at least two weeks according to World Health Organization).^16^ Further, additional cost reduction strategies, such as pooled PCR testing, or partnering with state or local governments for reduced-priced PCR or serological testing should be considered.^17^

Our study has several limitations. First, we did not include the additional costs (including tests and personnel) related to contact tracing. While this may affect the point estimates of annual testing costs, it is unlikely to vary by testing strategy. Second, we assumed low PCR and serological test result misclassification, and did not consider potential cost or epidemic implications of PCR false negatives^18^ (owing either to sampling collection, specimen handling, storage condition problems or low viral load) or antibody false positive^19^ (such that people think they are immune when they are not) especially for larger size universities under the condition of high SARS-CoV-2 prevalence/incidence ^20^, which may incorrectly inflate the estimated prevalence and cumulative incidence. On the other hand, as we consider frequent screening for PCR (e.g at least once a week for majority students in campus regardless of symptoms), this may result in a number of false positive cases (and associated costs for contact tracing) for larger universities under the condition of low SARS-CoV-2 prevalence/incidence ^21^. The overall utility of screening strategies may differ, however, by prevalence of disease among the population and the potential associated costs of false positives and negatives.^22^ Third, we assumed that the immune response remains throughout the entire school year (32 weeks). If duration of immune response is shorter than the whole school year (e.g. 16-20 weeks)^23^ and antibodies cannot be reliably detected after several weeks, the initial prevalence of seropositive university community members and associated cost saving may be lower than the range we have assumed here. We also assumed no regular PCR testing if the antibody test is positive, but there is on-going research for serology test validation which may influence the accurate assessment of true immunity and prevalence for individual and population.^24^ While some degree of immunity to COVID-19 after recovery is assumed, it remains an open question whether the presence of antibodies correlates with protection from disease and whether there is a specific antibody level that correlates with immunity. ^25^ Fourth, we did not quantify the savings to the individual, in terms of their time and opportunity cost, and potentially bearing the cost of the testing depending on the payer of the testing strategy. At the individual level, particularly when seropositive, cost-savings could be substantial. Finally, we did not consider the addition of antigen testing yet as part of a testing strategy given the relatively low sensitivity compared to the PCR test. Future work should incorporate the use of rapid antigen testing as a component of an on-campus testing strategy.

## Conclusion

To conclude, while the total testing cost is likely much lower if regular PCR testing is insourced compared to outsourced ($5 million vs $34 million), the inclusion of serological testing alongside regular PCR testing can reduce total test volume and achieve cost savings up to 20% compared to PCR testing alone. As we discuss when and how to phase in re-opening, these results may be useful to decision-makers in planning SARS-CoV-2 testing and considering an investment for mass screening of PCR and serological testing strategies under various capacities of academic institutions and epidemic conditions in the United States. Given the substantial financial investment required for regular PCR testing on university campuses, serological testing may be one tool to reduce total costs without compromising the epidemic containment strategy.

## Data Availability

Data consisted of model output and parameters from existing literature. Data available upon request from corresponding author.

